# Quantifying the role of naturally- and vaccine-derived neutralizing antibodies as a correlate of protection against COVID-19 variants

**DOI:** 10.1101/2021.05.31.21258018

**Authors:** Jamie A. Cohen, Robyn M. Stuart, Katherine Rosenfeld, Hil Lyons, Michael White, Cliff C. Kerr, Daniel J. Klein, Michael Famulare

**Affiliations:** Institute for Disease Modeling, Global Health Division, Bill & Melinda Gates Foundation, Seattle, USA; Department of Mathematical Sciences, University of Copenhagen, Copenhagen, Denmark; Department of Global Health, Institut Pasteur, Paris, France

## Abstract

The functional relationship between neutralizing antibodies (NAbs) and protection against SARS-CoV-2 infection and disease remains unclear. We jointly estimated protection against infection and disease progression following natural infection and vaccination from meta-study data. We find that NAbs are strongly correlated with prevention of infection and that any history of NAbs will stimulate immune memory to moderate disease progression. We also find that natural infection provides stronger protection than vaccination for the same level of NAbs, noting that infection itself, unlike vaccination, carries risk of morbidity and mortality, and that our most potent vaccines induce much higher NAb levels than natural infection. These results suggest that while sterilizing immunity may decay, we expect protection against severe disease to be robust over time and in the face of immune-evading variants.

## Introduction

As the COVID-19 pandemic has entered its second year, two trends have developed that are shaping ongoing transmission and disease dynamics. First, variants of concern that replicate more quickly and/or have the ability to evade existing immunity in individuals have emerged and spread globally. Second, COVID-19 vaccines are being rolled out, but coverage is highly heterogeneous. A nuanced understanding of immunity in individuals and populations is required to design policy that optimally allocates vaccines and implements non-pharmaceutical measures to counter the impact of emerging variants.

Immunologic memory, including neutralizing antibodies (NAbs), CD4+ and CD8+ T cells and memory B cells, is the basis for protection against infection and disease [1]. NAbs bind to viral proteins, blocking infection, while CD8+ T cells target virus cells, moderating disease severity. SARS-CoV-2 infection and vaccination induce a robust NAb response within individuals, followed by decay over time [2, 3]. An open question remains how a history of immune memory, from natural infections and/or vaccination, will protect against future risk of infection, onward transmission, and disease severity.

A known correlate of protection for SARS-CoV-2 and COVID-19 is urgently needed in order to assess population protection and expedite vaccine trials by enabling licensure based on immune readouts rather than large phase 3 trials [4]. Recent work from Khoury et al. [5] and Earle et al. [6] relates neutralizing antibodies to vaccine efficacy, showing that neutralization level is highly predictive of immune protection from symptomatic COVID-19, and that despite decaying immunity, protection from severe disease should be largely retained. However, this work did not consider immune protection from natural infection and did not disentangle the relationship between NAbs and protection against primary infection, symptomatic, and severe disease. Our analysis aims to fill these gaps and extend the existing work.

In our study, we model protective efficacy as a function of neutralizing antibodies (NAbs), informed by data from vaccine immunogenicity and efficacy trials as well as observational studies. Extending upon the methodology from Earle [6] and Khoury [5], we estimate efficacy for primary SARS-CoV-2 infection, symptomatic COVID-19, and severe disease jointly and calculate conditional efficacies for symptom- and severity-blocking given infection, revealing a direct model of NAbs as a correlate of protection. We additionally consider differences between the neutralizing antibodies generated from vaccines and natural infection while accounting for antibody waning between immunogenicity and vaccine efficacy studies. Finally, we describe a method for inferring protection against variants of concern over time.

## Results

### Efficacy against infection, symptomatic COVID-19, and severe disease

We find that neutralizing antibodies are strongly correlated with protection against SARS-CoV-2, symptomatic COVID-19, and severe disease (see Fig 1). In order to provide a 50 percent or higher reduction in the risk of symptomatic COVID-19, a vaccine would need to induce a NAb level at least one-tenth of the average convalescent level, and a one-third NAb level would be required to reduce the risk of infection by 50 percent or higher. While natural infection provides greater protection than vaccination for the same level of NAbs, all of the vaccines considered in this analysis meet the 50 percent risk reduction threshold and do not have the morbidity and mortality costs associated with COVID-19 infection. However, as will be discussed below, variants of concern challenge the efficacy of vaccines by reducing the neutralization levels and associated protection.

**Fig 1.**

Immunity model. We jointly estimated the relationship between NAbs and vaccine efficacy against infection, symptomatic, and severe disease using meta-study data. NAbs are normalized relative to human convalescent sera and central estimates are plotted. Panels show fitted efficacy against (A) infection, (B) symptomatic COVID-19, and (C) severe COVID-19 as a function of natural-and vaccine-derived NAbs. Shaded regions represent 95% credible intervals with *γ*_*k*_ fixed at it’s median.

Given the fitted marginal efficacies above, we inferred the conditional protection against symptomatic and severe disease for individuals with a breakthrough infection and with breakthrough symptomatic disease. We find that any history of immunity would provide some protection against symptomatic and severe COVID-19, with a floor of approximately 20 percent reduction in the risk of symptomatic COVID-19 and severe disease conditional on a breakthrough infection or disease. From that level, a percentage increase in NAbs would result in 3.8 percent (0.12, 11.75) reduction in risk against symptomatic disease a 7.9 percent (0.22, 26.15) reduction in the risk of severe COVID-19, conditional on a breakthrough infection and breakthrough disease respectively. While NAbs are correlated with protection against COVID-19, there may be other immune mechanisms, such as T-cell response, that provide protection against symptomatic and severe disease [7]. These results suggest that even as antibodies wane and become insufficient to protect against infection, some immunity to symptomatic and severe disease will remain.

### Efficacy over time

We use this fitted model to infer the extent to which protective efficacy will decay as NAbs wane and in the face of immune-evading variants. We compared immunity to a future alpha or delta variant challenge based on prior immunity from natural infection, and 1- and 2-doses of the Pfizer and AstraZeneca vaccines. For each of these, we assume the same antibody waning kinetics but different peak NAb levels and different neutralization capacity against a delta variant challenge, see Supplemental Appendix for details.

With respect to protection against infection, immunity from either 1-dose of the Pfizer vaccine or both 1- and 2-doses of the AstraZeneca vaccine provides less than 50% protection with a delta variant almost immediately and decays from there, see Fig 2. Immunity from a prior natural infection would fall below 50% protective efficacy within one year. In contrast, two doses of Pfizer is able to provide over 80% protection against infection for over one year.

**Fig 2.**
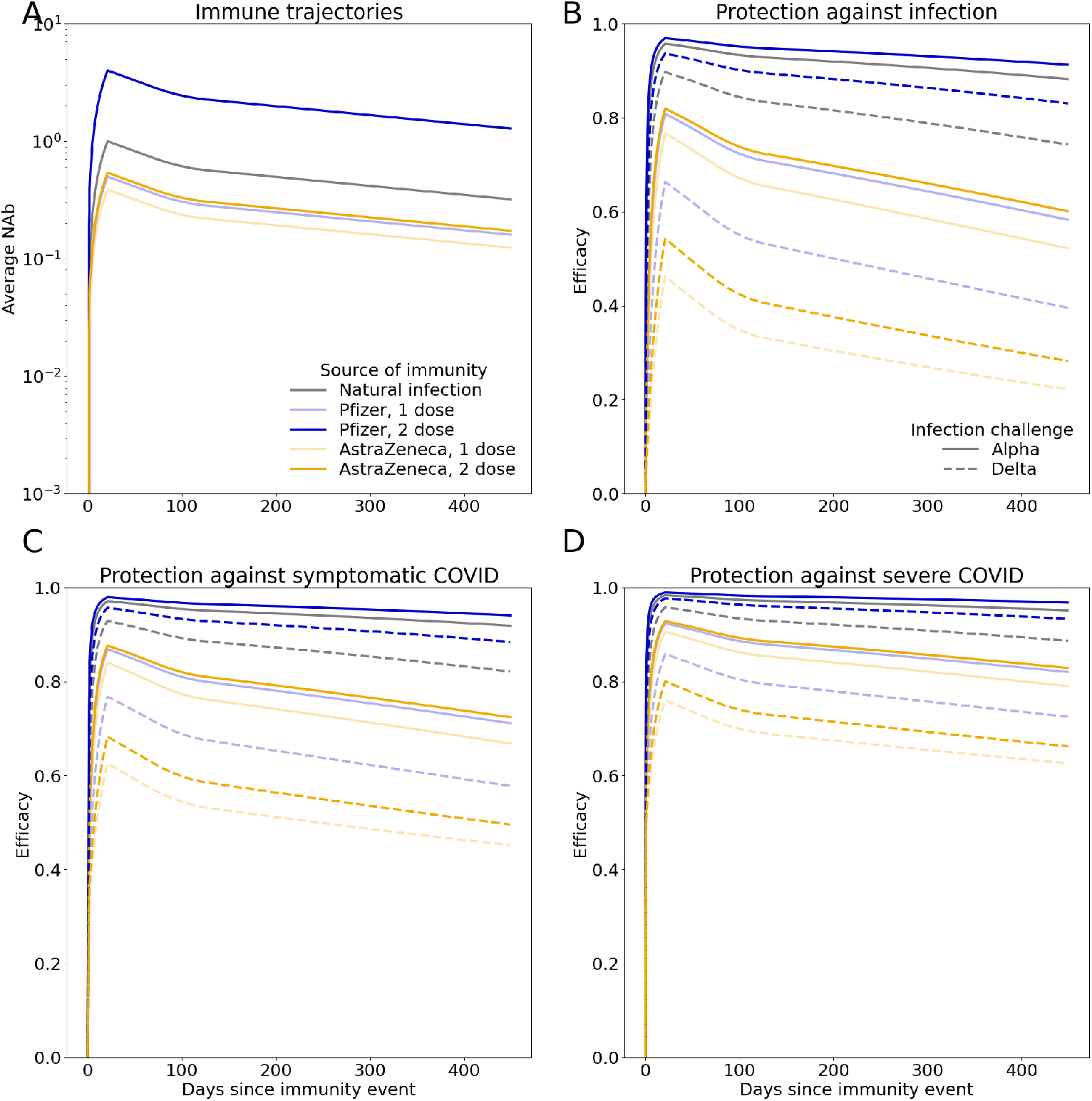
Inferred efficacy over time. We use a model of antibody waning to estimate how much immune protection will wane over time given different sources of immunity and in the face of an immune evading variant. Panel (A) shows immune trajectories over time, (B)-(D) show the efficacy against infection, symptomatic, and severe COVID, respectively, over time. The solid lines represent risk reduction against an alpha variant and dashed lines represent risk reduction against a delta variant. Color indicates source of immunity.

Despite decaying protection against infection, protection against severe disease is notably retained for all immune trajectories considered, with even a single dose of AstraZeneca reducing the risk of severe disease caused by delta by over 60% a year after vaccination. These results underscore the potency of vaccines against severe disease. They also reveal how breakthrough infections can be expected with increasingly immune evading variants, but protection against severe disease will be robust.

## Discussion

As efficacy results from COVID-19 vaccine trials began to appear in late 2020, there was new optimism that the worst of the pandemic may have passed [8]. However, this optimism was quickly curtailed by the simultaneous discovery of mutations of SARS-CoV-2 that led to extreme epidemic resurgences around the world, including in countries that had already experienced high epidemic burdens. These two developments implied a considerable shift in the COVID-19 epidemic landscape, and called for new approaches to help guide responses.

As evidence builds that vaccine efficacy varies considerably across vaccines and variants [9–12], and while questions around the duration of protection afforded by both vaccination and infection linger [13–18], there is an increasing need to identify a correlate of protection for SARS-CoV-2 and for models that can account for these factors. Despite the rapid pace of COVID-19 science, key immune dynamics, including immune memory over time, are missing from virtually all models that are increasingly relied upon to inform policy [19]. This study extends recent work estimating the relationship between NAbs and protection against infection and progression to symptomatic and severe disease. The results of this study can be used independently or integrated into models of SARS-CoV-2 and COVID-19.

Our results show that while NAbs induced by natural infection or vaccination wane and individuals may lose sterilizing immune protection, immune memory is likely to be retained long-term to provide significant protection against severe disease, even in the face of immune-evading variants. This suggests that neutralizing titers play a large role in preventing infection, but that other immunologic factors may play a more dominant role in controlling infection once it occurs.

Our modeling approach relies on estimating a relationship between NAbs and protection against infection, symptomatic disease, and severe disease, and the data used to establish these estimates are scarce and uncertain, especially for low levels of NAbs. While a full individual-level model would be ideal, we relied upon published cohort averages and tried to account for variation and heterogeneity between studies using study-level random effects. We also assume that the antibody kinetics are identical for vaccine- and naturally-derived NAbs. As more longitudinal immune studies emerge, we will have the opportunity to test and refine this hypothesis. We do not specifically model cellular immune responses, although they are likely to also influence disease symptomaticity and severity and to have different kinetic profiles than antibodies [7, 20].

## Methods

### Data Extraction and Estimation

In order to map NAb level to protective efficacy, we extracted cohort estimates from vaccine immunogenicity and efficacy trials as well as data on reinfection (see Supplemental Appendix for details). In the absence of standardized assays to measure NAbs, normalization against a convalescent serum standard has been suggested as a method for providing greater comparability between results from different assays [21]. In order to compare the immunogenicity data with the efficacy endpoints, we accounted for waning that may have occurred across the timescales reported. We re-normalize the average NAb for each of the cohorts using an adaptation of the antibody kinetics functional form described in Khoury et al. [5] fit to cohorts of hospitalized patients and healthcare workers followed-up for eleven months after COVID-19 symptom onset [22]. See Supplemental Appendix for additional details.

We also adjusted the reported neutralization level in settings where variants of concern were circulating at the time of efficacy endpoints based upon reported neutralization in convalescent and vaccine sera (see Supplemental Appendix).

### Immune Model

We modeled three types of immunity: protection against infection, symptomatic disease, and severe disease. Once we consider the impact of waning immunity in the re-normalization of cohort-average NAbs, we find that it is challenging to fit protection against infection with a single curve for both vaccine- and natural infection-derived immunity (see Supplemental Appendix). Therefore, we fit separate functions for these two sources of immunity, which can be supported by the role of nucleocapsid-specific antibodies which are missing from some vaccines and may mechanistically explain why natural NAbs are more effective against infection [23].

We jointly estimated the relationship between NAbs and protective efficacy against infection, symptoms, and severe disease with study-specific random effects. VE_symp|inf,*r*_ and VE_sev|symp,*r*_ are unobserved, and we model them through the marginal efficacy against symptomatic and severe disease (Eqs 1 and 2).

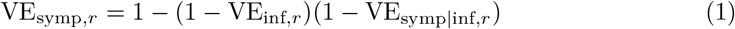

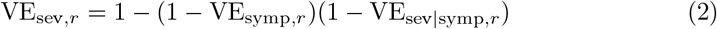

We split vaccine efficacy into conditional parts to match the stages of the infection process and assumed both the efficacy against infection and the conditional efficacy against symptoms and severe disease are logit-log. The *α* and *β* parameters capture the intercept and slope in each equation, respectively (see Eqs 3, 4, 5). Vaccine efficacy against infection, VE_inf_, is the first stage that modulates the probability of infection given exposure. For people who get infected, symptomaticity is modulated by the conditional vaccine efficacy given breakthrough infection, VE_symp| inf_, and similarly severity is modulated by the conditional vaccine efficacy given a breakthrough symptomatic infection VE_sev|symp_.

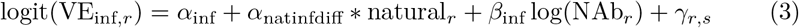

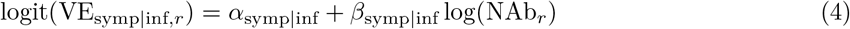

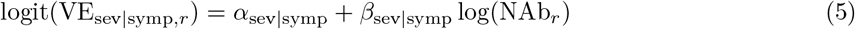

where NAb_*r*_ represents the average level of neutralizing antibodies across participants in record *r*, “natural” is a dummy variable that is equal to 1 when record *r* is immunity from natural infection and equal to 0 when record *r* is immunity from vaccination, and *γ*_r,s_ is the random effect from study *s* associated with record *r* (some studies have multiple associated records).

For studies which reported efficacy against variants of concern, as part of the re-normalizing computational procedure, NAbs were randomly shifted by a normally-distributed scaling factor with a mean and standard deviation based upon titer shifts reported in the literature [14, 24–28, 28]. That is, the results marginalize over uncertainty in the NAb titer shift. Additional details are provided in the appendix.

We estimated a Bayesian posterior for parameters in Eqs 3 - 5 with a Hamiltonian Monte Carlo method fit in Stan based upon the likelihood (Eqs 6 - 8). 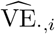 denotes vaccine efficacy estimators arising from the studies, that is, the data. The likelihood uses the standard errors as reported by the studies adapted to a log-scale as a reasonable proxy for the true standard deviation, one that incorporates study sample size; details may be found in the appendix.

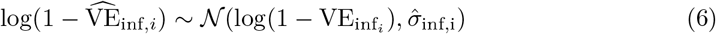

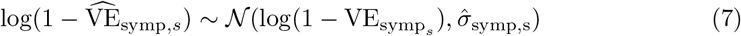

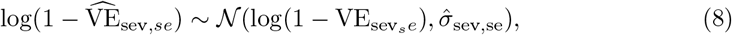

for *k* in *K* unique study cohorts, *i* in *I* infection records, *s* in *S* symptomatic disease records, and *se* in *SE* severe disease records.

In the above equations, *σ*_*γ*_ is the standard deviation of the study random effects, *γ*_*s*_ represents the study-specific random effect *s* in *S* unique studies, and 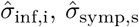 and 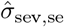 represent the log-scale standard deviation of outcomes of protection against infection, symptomatic and severe disease. Further computational details may be found in the Supplemental Appendix.

## Data Availability

All data and code are freely available.

https://github.com/amath-idm/COVID-Immune-Modeling

## Acknowledgments

We thank Rafael Núñez, Bradley Wagner, Stewart Chang, Amanda Izzo, and Jennifer Schripsema for their thoughtful review and guidance, as well as David Khoury, Miles Davenport, George Kassiotis, and David Bauer for their helpful comments.

