## Supplemental Appendix for "Quantifying the role of naturally- and vaccine-derived neutralizing antibodies as a correlate of protection against COVID-19 variants"

### Data extraction and estimation

In the NAb re-normalization procedure, we used a model of immune waning to account for any decay in antibodies that may have occured between the time of the antibody assay collection and vaccine efficacy endpoints. To do so, we assume waning follows a 2-part exponential decay and fit the half-life and duration parameters to cohorts of French and Irish hospitalized patients and healthcare workers followed for up to eleven months after COVID-19 symptom onset [1]. Relative to the waning model used by Khoury et al., our model suggests both shorter initial decay of NAbs followed by a steeper long-term decay rate, see Fig 1.

### Estimation procedure

We have implemented study random effects in our infection regression model, which we assumed to be normally distribution with a mean of 0 and a standard deviation that is Cauchy distributed with a flat prior,

$$\gamma_s \sim \mathcal{N}(0, \sigma_\gamma) \tag{1}$$

$$\sigma_\gamma \sim Cauchy(0, 1). \tag{2}$$

In order to compute titer shifts for efficacy against variants of concern, NAbs were randomly shifted by a normally-distributed scaling factor,

$$\text{titer shift}_s \sim \mathcal{N}(\text{titer shift mean}_s, \text{titer shift SD}_s) \tag{3}$$

with a mean and standard deviation based upon titer shifts reported in the literature, see Table 1.

| Variant | Wild-Type | Pfizer | Moderna | AstraZeneca |
|---|---|---|---|---|
| Alpha (B.1.1.7) | 1.8 (0.41) | 1.8 (0.41) | 1.8 (0.41) | – |
| Beta (B.1.351) | 13.3 (2.17) | 10.3 (2.9) | 12.4 (2.85) | – |
| Gamma (P.1) | 8.66 (1.05) | – | – | – |
| Delta (B.1.617.2) | – | 2.19 (0.05) | – | 4.01 (0.32) |

**Table 1.** Variant neutralization with wild-type and vaccine-sera. Values refer to the fold of reduction in neutralization with each variant relative to wild-type infection or vaccine source, standard deviation reported in parentheses.

Model fitting was performed in R using Stan. We report the mean and standard deviation of the model parameters in Table 2.

We also considered a single curve for naturally- and vaccine-derived NAbs, but found that this form systematically under-estimated the degree of protection found in the convalescent reinfection studies, see Fig 2.

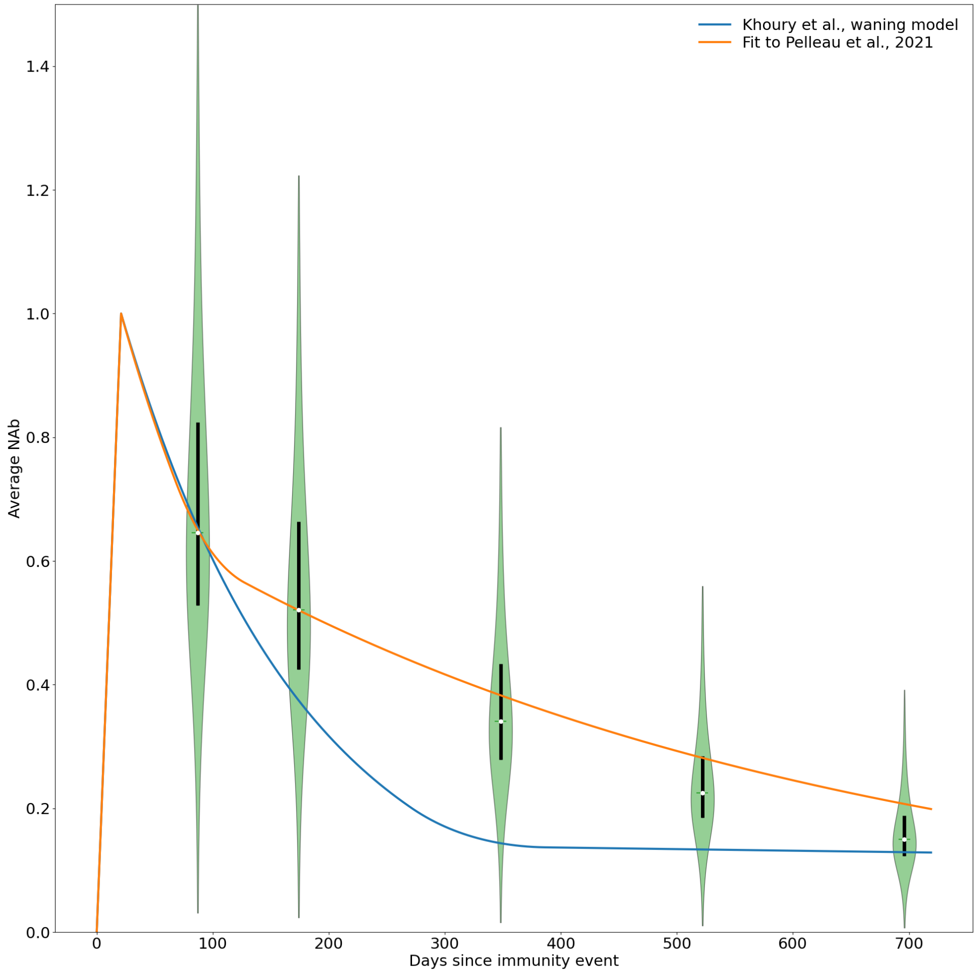


**Fig 1.** Comparison of waning immunity models.

| | Mean | Standard Deviation |
|---|---|---|
| $\alpha_{\text{inf}}$ | 1.08 | 0.226 |
| $\alpha_{\text{natinfdiff}}$ | 1.0 | 0.403 |
| $\beta_{\text{inf}}$ | 0.967 | 0.055 |
| $\alpha_{\text{symp}\mid\text{inf}}$ | -0.739 | 0.187 |
| $\beta_{\text{symp}\mid\text{inf}}$ | 0.038 | 0.0317 |
| $\alpha_{\text{sev}\mid\text{symp}}$ | -0.0143 | 0.311 |
| $\beta_{\text{sev}\mid\text{symp}}$ | 0.0799 | 0.078 |
| $\sigma_{\gamma}$ | 0.639 | 0.177 |

**Table 2.** Fitted parameter values based upon HMC algorithm with 30,000 iterations across 5 chains.

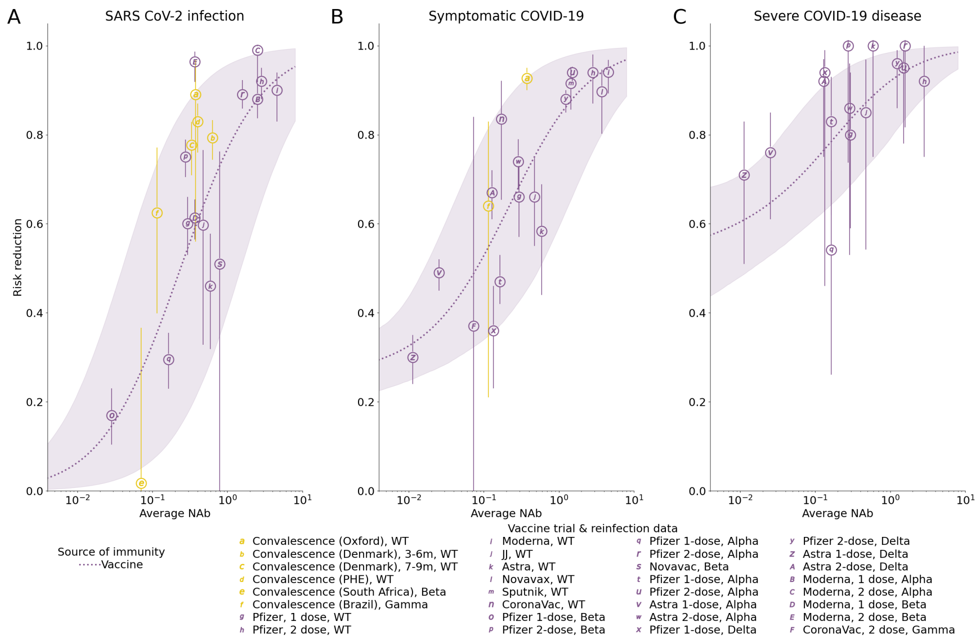


**Fig 2.** Joint estimation with one curve for both natural- and vaccinal-immunity.
